# Development and validation of the inflammatory bowel disease objective disability index (IBDODI)

**DOI:** 10.1101/2023.07.11.23292511

**Authors:** Luigi Melcarne, Eduard Brunet, Albert Villoria, Mercedes Vergara, Anna Puy, Laura Llovet, Belen García-Sague, Oliver Valero, Xavier Calvet

**Affiliations:** Unidad Enfermedad Inflamatoria Intestinal – Servicio Aparato Digestivo - Parc Taulí Hospital Universitari. Institut d’Investigació i Innovació Parc Taulí (I3PT-CERCA). Sabadell, Spain; Departament de Medicina. Universitat Autònoma de Barcelona. Sabadell, Spain; CIBERehd, Instituto de Salud Carlos III, Madrid, Spain; Servei d’Estadística Aplicada - Universitat Autònoma de Barcelona. Cerdanyola, Spain

## Abstract

**Introduction:** Many scores aiming to measure Inflammatory bowel disease (IBD)-related disability have been validated. However, they rely mostly on patients’ subjective perceptions, and may, therefore, be biased by them. To obtain a more objective measure of disability, the present study aimed to develop and validate an IBD-related disability index based on objective and measurable variables.

**Methods:** Answering an online survey, IBD patients reported their officially recognised degree of general and work disability plus a number of objective parameters for its assessment. Responses were randomly allocated to one of two datasets: a) training or b) validation. Multiple logistic regression tests were performed in the training set. Variables that were statistically or clinically significant were used to model two scores: The Unweighted IBD Objective Disability Index (U-IBDODI), where all variables had the same weight, and the Weighted IBD Objective Disability Index (W-IBDODI), in which variables were weighted according to the regression results. Both scores were subsequently validated.

**Results:** The analysis included 930 valid questionnaires. Patients’ mean age was 41±11, 642 (65.4%) were women, and 582 (59.3%) had Crohn’s Disease. The training dataset included 665 surveys. In the validation set (n=265), U-IBDODI mean values were 3.7 +/-1.3 for patients with work disability and 2.3+/-1.4 for patients without (p<0.001). U-IBDODI AUROCs for predicting work and general disability were 0.839 and 0.675 respectively. The corresponding W-IBDODI values were 10.9 +/-3.7 and 6.9+/-3.7 (p<0.001), 0.837 and 0.606.

**Discussion:** Both U-IBDODI and W-IBDODI scores are easy-to-use, objective and valid tools for measuring work disability in IBD patients.

**DISCLOSURE STATEMENT:** LM has served as a speaker and consultant for MSD, Abbvie, Janssen, Takeda, Tillotts, Kern, Chiesi. AV has served as a speaker and consultant for MSD and Abbvie. EB has served as a speaker and consultant for Jansen, Kern and Chiesi. XC has received grants for research from Janssen, Kern, Abbott, MSD, Pfizer, Galapagos and Vifor, and fees for advisory board services from Janssen, Abbvie, MSD, Takeda and Vifor. He has also given lectures for Janssen, Abbvie, MSD, Takeda, Shire and Allergan.

**AUTHORS’ CONTRIBUTIONS:** LM and XC designed the study, analyzed the data and wrote the manuscript. OV analyzed data. LM, XC, EB, AV, MV, AP, LL, OV critically reviewed the text and provided important intellectual content. All authors definitively approved the submitted version.

**FUNDING:** The project won the VII "Antonio Obrador" grant from GETECCU (grupo español de trabajo en enfermedad de Crohn y Colitis ulcerosa - Spanish working group on Crohn’s disease and Ulcerative Colitis

## Introduction

Disability is defined as the partial or total inability to perform social roles, such as work activity, in a manner consistent with norms or expectations. Inflammatory bowel diseases (IBD) are a frequent cause of disability [1]. IBD – either Crohn’s disease (CD) or ulcerative colitis (UC) – are immunomediated diseases affecting the digestive tract, characterised by a chronic clinical course with phases of remission and flares of activity. The prevalence of IBD in Western countries is high and seems to be increasing progressively worldwide [2–4]. Persistent inflammatory activity or sequelae derived from abdominal surgery frequently lead to cessation of work activity and disability, often permanent, especially in the subgroup of patients with severe disease [5]. The effect is especially important given the fact that IBD is more prevalent in young, socially and occupationally active individuals [6]. Multiple factors are involved in IBD-related disability such as the severity and duration of the disease, the presence and severity of associated diseases, hospital admissions, the use of biological drugs and abdominal surgeries [6].

The disability rate varies considerably from 1.3% to 50% depending on the study [1,6–12]. This great variability may be caused in part by the different definitions of work disability and the variety of the tools used to measure it. In Spain, approximately one in 25 (4.1%) IBD patients receive a work disability pension [5].

Several scores of general [13–15] and work disability [16–20] have been validated. Specifically, our group validated a self-reported work disability questionnaire for Crohn’s disease that initially contained 16 items and was subsequently reformulated and validated in a 10-item format [16]. This self-reported disability questionnaire was later validated for ulcerative colitis patients [17]. Self-reported questionnaires, however, may be heavily biased by patients’ subjective perceptions. In addition, it is important to distinguish between temporary disability –for example, during a flare of the disease– and permanent or long-term disability secondary to surgical sequelae or refractory persistent disease. To our knowledge, no questionnaires have been developed to date to objectively evaluate permanent work disability. A reliable, thorough-going questionnaire of this kind would be particularly useful for the health authorities and public bodies responsible for evaluating and granting work disability benefits. The Spanish health authorities measure disability in two ways. General disability [21] is evaluated at regional level, while work disability in active workers is evaluated by the national Social Security service. A previous study by our group showed that the awarding of work disability pensions to applicants varied markedly depending on the evaluating tribunal [22]. So, tools for measuring permanent disability based on objective parameters might help to increase impartiality and fairness in the granting of disability benefits.

This study aims to develop and validate an index of disability for patients with IBD based on a set of objective parameters.

## Methods

### Subjects

Adult patients with IBD diagnosed at least six months previously and legally resident in Spain were invited to participate in the study by completing an online survey. Among those who agreed to take part, those with repeated or incomplete surveys (i.e., with fewer than 50% valid responses) and those who did not meet the inclusion requirements (primarily those aged under 18) were excluded.

### Survey development

An online survey was performed using the SurveyMonkey© platform performed between January 2019 and June 2019. Based on the experience and results of an earlier population study published by our research group [5] the survey was designed to be answered by patients without the need for health professionals’ support.

The survey link was posted on the websites of IBD patients’ associations such as *ACCU España* and *ACCU Catalunya*. These associations also publicised the survey among their members on social media.

Objective and measurable parameters were chosen for the survey after reviewing the data evaluated in our previous study [5]. Patients were asked about their age, sex, years of disease evolution, type of inflammatory bowel disease (CD or UC), extension, mean number of depositions over the last six months, fecal incontinence, treatment, number of radiological or endoscopic examinations, days of hospitalisation and days off work during the last year, number of previous surgeries secondary to IBD, ostomies, psychiatric disorders diagnosed and psychoactive drugs prescribed. Incontinence was measured by the Wexner score and classified as “no incontinence” (Wexner = 0), “mild” (Wexner ≥1 ≤8) and moderate/severe (Wexner ≥ 9). Patients also reported their officially acknowledged degree of disability. In accordance with the Spanish social security structure, the survey asked about both work disability and general disability. Work disability was evaluated only in active or past workers, while general disability could be evaluated in all patients. The degree of patients’ general disability is measured as a percentage, and is then categorised into four groups: no disability (0%), low (1-33%), moderate (34-55%) and severe disability (> 55%). Each of these grades receives a different level of social support, and individuals with a severe degree of disability and a low income receive economic aid. Active or past workers reported whether they were granted a work disability pension and also recorded their officially acknowledged degree of general disability.

Patients were also asked to complete the following questionnaires:

- SCCAI (Simple Clinical Colitis Activity Index) for patients with ulcerative colitis [23] and the Harvey-Bradshaw Index for patients with Crohn’s disease, in order to evaluate disease activity [24].
- A validated self-reported questionnaire on disability in Crohn’s disease and ulcerative colitis [16,17] and the international index of disability in inflammatory bowel disease (IBD-DI), to evaluate global and work disability in IBD[25][13,14,25].
- EuroQol 5D3L a self-administered generic questionnaire assessing quality of life [26].
- IBDQ-9 (Inflammatory Bowel Disease Questionnaire 9) quality of life questionnaire in inflammatory bowel disease [27]. This is a shortened and validated version of the IBDQ-36 quality of life questionnaire for patients with IBD [28].
- WPAI (work productivity and activity impairment questionnaire) [29] to evaluate work complaints due to IBD.

Of these surveys, the completion of the disease activity questionnaire and the self-reported questionnaire on disability was considered mandatory.

### Statistical methods

The completed questionnaires were randomly assigned to either the training dataset (67% of patients) or the validation dataset (33%).

#### Bivariate analysis

In the training sample, a bivariate test was carried out to determine the variables related to three different outcomes: ’work disability’, ’general disability’ and ’degree of general disability’.

‘Work disability’ was defined in cases of patients of working age receiving disability benefit. Questionnaires completed by students, housemakers and retirees were not included in the work disability analysis. General disability (yes/no) was defined in cases with any recognised degree of disability. Finally, general disability was also evaluated as a categorical variable with four categories (no, mild, moderate and severe) according to the criteria mentioned above. Qualitative variables were compared using the Chi-Squared Test, Fisher’s Exact Test or Likelihood Ratios.

For quantitative variables, the application conditions of the different tests (Shapiro-Wilk normality tests and Levene’s Homogeneity of variance tests) were analysed. The appropriate test was used based on compliance with the application criteria (T-test/ANOVA or Mann-Whitney-Wilcoxon Test/Kruskal Wallis Test).

#### Multivariate analysis

Three multivariate logistic regression models were performed, one for each response variable (’work disability’, ’general disability’ and ’degree of general disability’).

Variables with a *p*-value less than 0.1 in the bivariate analysis were included in each of the multivariate logistic regression models. For the variable ’degree of general disability’ an ordinal logistic regression model was used using the four categories of disability defined above. Three final models were obtained after eliminating non-statistically significant variables and maintaining those that were clinically relevant. The results were represented graphically with ROC curves, and the value of the area under the curve (AUROC) was calculated.

#### Modelling the global disability score

A single general model was designed to predict disability. The variables that were present in at least two of the logistic regression models were selected to construct a final score based on objective variables that could be useful for predicting both work and general disability in IBD. Variables that did not fulfil the above conditions but were considered relevant by the authors according to the literature were also included in the model.

Two different scoring systems were tested for this final model. In the first, the score was calculated by assigning the same weight to each risk factor, and in the second, a different weight was assigned to each predictive variable based on the odds ratio values of the individual models.

#### Validation

The predictive capacity of the two scoring systems was evaluated in both the training and the validation sets. It was represented graphically using the ROC curves, and the AUROC was calculated for each scoring system.

All statistical analyses were performed with the software SAS v9.4, SAS Institute Inc., Cary, NC, USA. A p value of 0.05 was considered significant.

#### Sample size calculation

A convenience sample approach was used, including all the valid surveys in each of the analyses.

### Ethical issues

The study was approved by the ethics review board of Parc Taulí University Hospital (2019/523). The study was registered as clinical trial NCT03872726.

Patients gave on-line informed consent to participate before completing the questionnaire. All study procedures were performed in accordance with the Helsinki Declaration [30]. Personal data were managed in accordance with the General regulation for data protection of the European Union (GDPR-EU Regulation 2016/679).

The anonymity of the patients was guaranteed. The study was performed and reported in accordance with the TRIPOD statement recommendations [31]. The TRIPOD checklist is available in supplementary table 1.

## Results

### Study population

A total of 1074 patients answered the survey, and 930 questionnaires were considered valid for the analysis (Fig 1). Regarding demographic data, 642 respondents (65.4%) were women, mean age was 41 (± 11) and 582 patients (59.3%) had CD. Half of the patients were on immunomodulatory or biological treatment. The most frequent biological treatments were infliximab (37%) and adalimumab (37%) (Table 1). A flowchart of the study is shown in figure 1. Eight hundred (86%) patients were classified as “workers”; of these, 532 (58.7%) were actively working, 103 (11.4%) were unemployed, 91 (10%) were on temporary sick leave and 74 (8.2%) had a work-disability pension.

**Figure 1:**
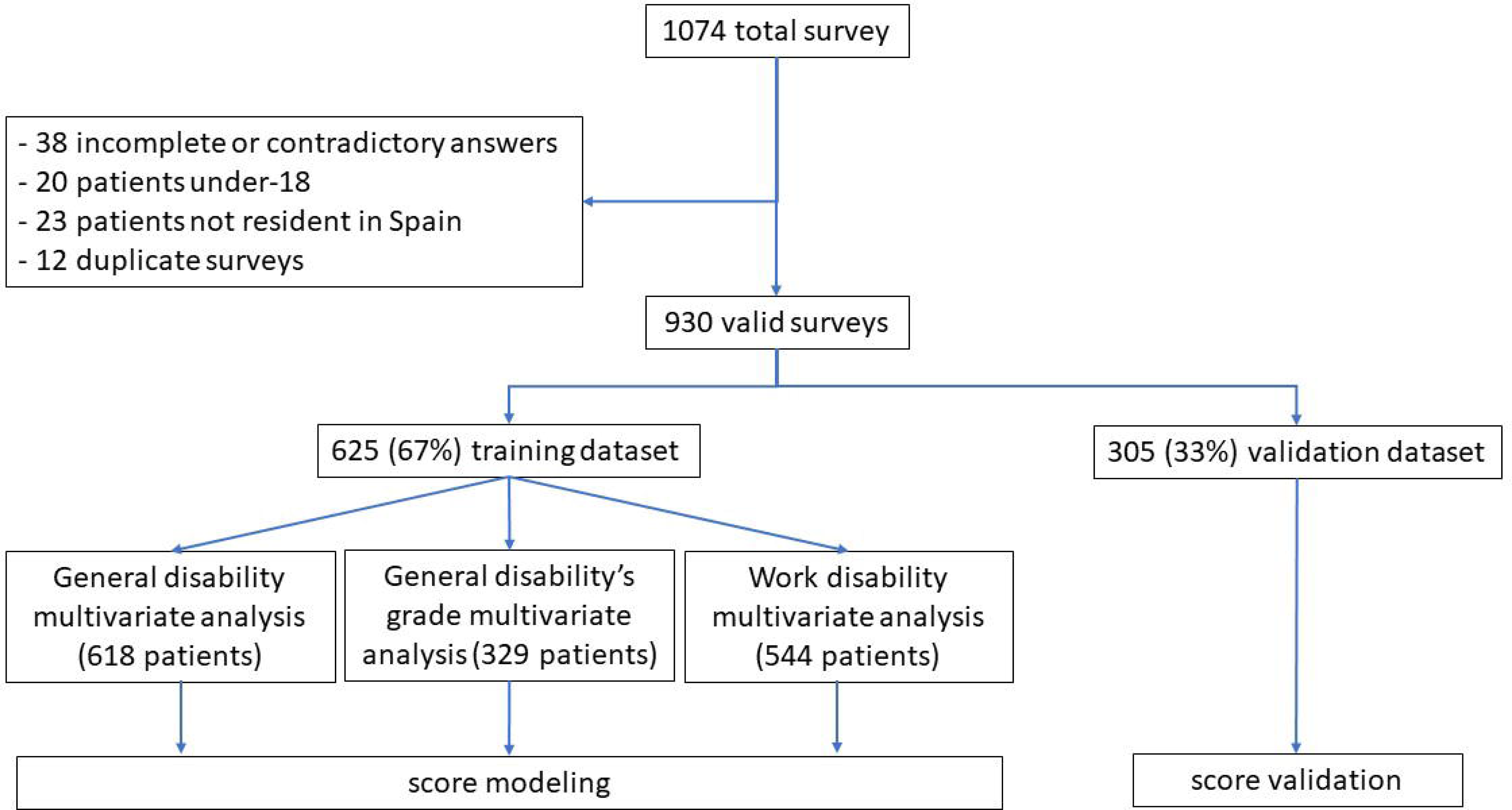
Study flowchart

**Table 1:**
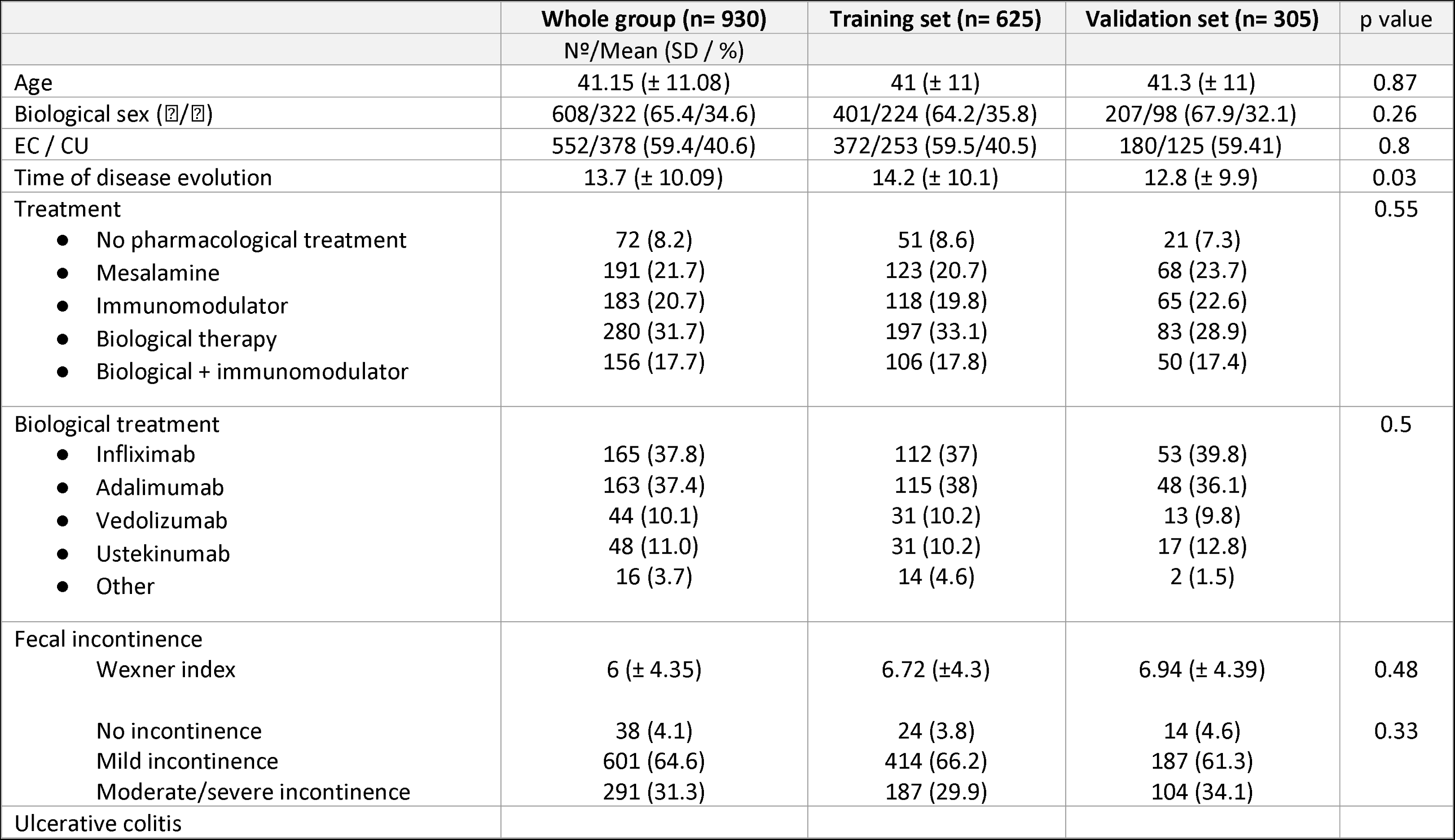
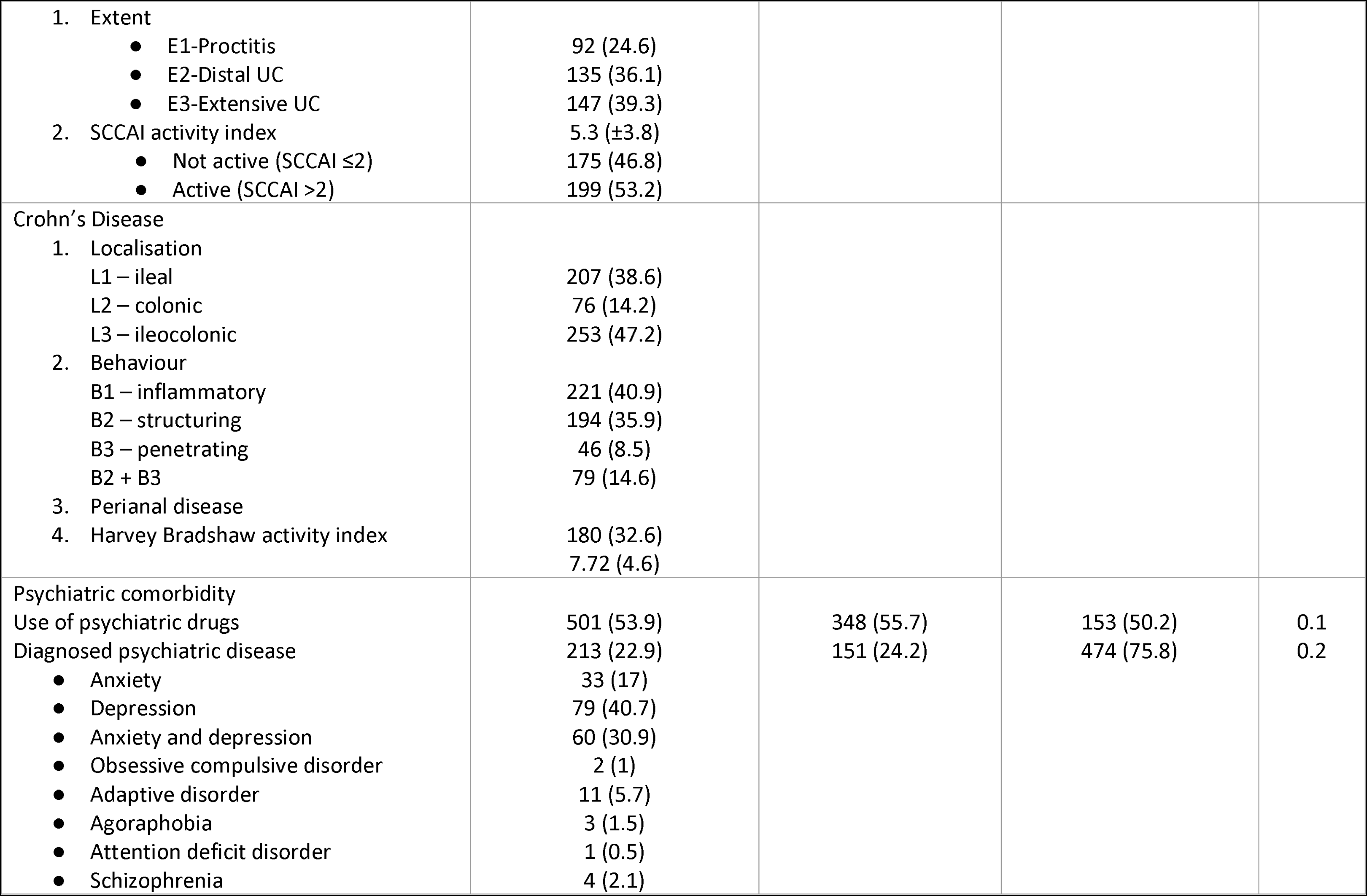
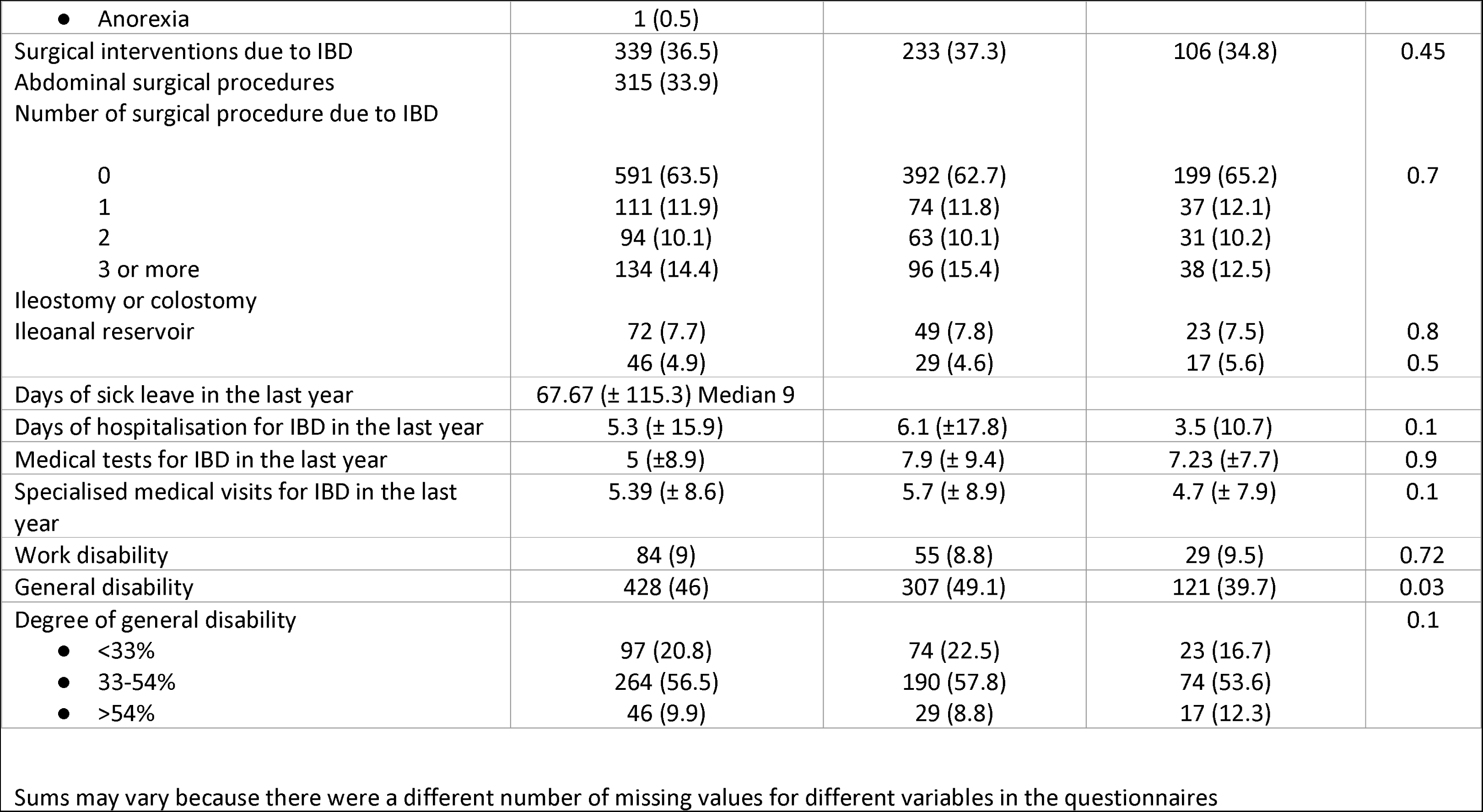
Baseline characteristics of the cohort

|  | Whole group (n= 930)<br>Nº/Mean (SD / %) | Training set (n= 625) | Validation set (n= 305) | p value |
| --- | --- | --- | --- | --- |
| Age | 41.15 (± 11.08) | 41 (± 11) | 41.3 (± 11) | 0.87 |
| Biological sex (♀/♂) | 608/322 (65.4/34.6) | 401/224 (64.2/35.8) | 207/98 (67.9/32.1) | 0.26 |
| EC / CU | 552/378 (59.4/40.6) | 372/253 (59.5/40.5) | 180/125 (59.41) | 0.8 |
| Time of disease evolution | 13.7 (± 10.09) | 14.2 (± 10.1) | 12.8 (± 9.9) | 0.03 |
| Treatment |  |  |  | 0.55 |
| • No pharmacological treatment | 72 (8.2) | 51 (8.6) | 21 (7.3) |  |
| • Mesalamine | 191 (21.7) | 123 (20.7) | 68 (23.7) |  |
| • Immunomodulator | 183 (20.7) | 118 (19.8) | 65 (22.6) |  |
| • Biological therapy | 280 (31.7) | 197 (33.1) | 83 (28.9) |  |
| • Biological + immunomodulator | 156 (17.7) | 106 (17.8) | 50 (17.4) |  |
| Biological treatment |  |  |  | 0.5 |
| • Infliximab | 165 (37.8) | 112 (37) | 53 (39.8) |  |
| • Adalimumab | 163 (37.4) | 115 (38) | 48 (36.1) |  |
| • Vedolizumab | 44 (10.1) | 31 (10.2) | 13 (9.8) |  |
| • Ustekinumab | 48 (11.0) | 31 (10.2) | 17 (12.8) |  |
| • Other | 16 (3.7) | 14 (4.6) | 2 (1.5) |  |
| Fecal incontinence |  |  |  |  |
| Wexner index | 6 (± 4.35) | 6.72 (±4.3) | 6.94 (± 4.39) | 0.48 |
| No incontinence | 38 (4.1) | 24 (3.8) | 14 (4.6) | 0.33 |
| Mild incontinence | 601 (64.6) | 414 (66.2) | 187 (61.3) |  |
| Moderate/severe incontinence | 291 (31.3) | 187 (29.9) | 104 (34.1) |  |
| Ulcerative colitis |  |  |  |  |
| 1. Extent |  |  |  |  |
| • E1-Proctitis | 92 (24.6) |  |  |  |
| • E2-Distal UC | 135 (36.1) |  |  |  |
| • E3-Extensive UC | 147 (39.3) |  |  |  |
| 2. SCCAI activity index | 5.3 (±3.8) |  |  |  |
| • Not active (SCCAI ≤2) | 175 (46.8) |  |  |  |
| • Active (SCCAI >2) | 199 (53.2) |  |  |  |
| Crohn's Disease |  |  |  |  |
| 1. Localisation |  |  |  |  |
| L1 – ileal | 207 (38.6) |  |  |  |
| L2 – colonic | 76 (14.2) |  |  |  |
| L3 – ileocolonic | 253 (47.2) |  |  |  |
| 2. Behaviour |  |  |  |  |
| B1 – inflammatory | 221 (40.9) |  |  |  |
| B2 – structuring | 194 (35.9) |  |  |  |
| B3 – penetrating | 46 (8.5) |  |  |  |
| B2 + B3 | 79 (14.6) |  |  |  |
| 3. Perianal disease |  |  |  |  |
| 4. Harvey Bradshaw activity index | 180 (32.6) |  |  |  |
|  | 7.72 (4.6) |  |  |  |
| Psychiatric comorbidity |  |  |  |  |
| Use of psychiatric drugs | 501 (53.9) | 348 (55.7) | 153 (50.2) | 0.1 |
| Diagnosed psychiatric disease | 213 (22.9) | 151 (24.2) | 474 (75.8) | 0.2 |
| • Anxiety | 33 (17) |  |  |  |
| • Depression | 79 (40.7) |  |  |  |
| • Anxiety and depression | 60 (30.9) |  |  |  |
| • Obsessive compulsive disorder | 2 (1) |  |  |  |
| • Adaptive disorder | 11 (5.7) |  |  |  |
| • Agoraphobia | 3 (1.5) |  |  |  |
| • Attention deficit disorder | 1 (0.5) |  |  |  |
| • Schizophrenia | 4 (2.1) |  |  |  |
| • Anorexia | 1 (0.5) |  |  |  |
| Surgical interventions due to IBD | 339 (36.5) | 233 (37.3) | 106 (34.8) | 0.45 |
| Abdominal surgical procedures | 315 (33.9) |  |  |  |
| Number of surgical procedure due to IBD |  |  |  |  |
| 0 | 591 (63.5) | 392 (62.7) | 199 (65.2) | 0.7 |
| 1 | 111 (11.9) | 74 (11.8) | 37 (12.1) |  |
| 2 | 94 (10.1) | 63 (10.1) | 31 (10.2) |  |
| 3 or more | 134 (14.4) | 96 (15.4) | 38 (12.5) |  |
| Ileostomy or colostomy |  |  |  |  |
| Ileoanal reservoir | 72 (7.7) | 49 (7.8) | 23 (7.5) | 0.8 |
|  | 46 (4.9) | 29 (4.6) | 17 (5.6) | 0.5 |
| Days of sick leave in the last year | 67.67 (± 115.3) Median 9 |  |  |  |
| Days of hospitalisation for IBD in the last year | 5.3 (± 15.9) | 6.1 (±17.8) | 3.5 (10.7) | 0.1 |
| Medical tests for IBD in the last year | 5 (±8.9) | 7.9 (± 9.4) | 7.23 (±7.7) | 0.9 |
| Specialised medical visits for IBD in the last year | 5.39 (± 8.6) | 5.7 (± 8.9) | 4.7 (± 7.9) | 0.1 |
| Work disability | 84 (9) | 55 (8.8) | 29 (9.5) | 0.72 |
| General disability | 428 (46) | 307 (49.1) | 121 (39.7) | 0.03 |
| Degree of general disability |  |  |  | 0.1 |
| • <33% | 97 (20.8) | 74 (22.5) | 23 (16.7) |  |
| • 33-54% | 264 (56.5) | 190 (57.8) | 74 (53.6) |  |
| • >54% | 46 (9.9) | 29 (8.8) | 17 (12.3) |  |

Sums may vary because there were a different number of missing values for different variables in the questionnaires

Regarding general disability, 428 patients (45.9%) had an officially recognised general disability degree, which was moderate in 56.5% of cases (Table 1).

### Baseline homogeneity analysis

Six hundred and twenty-five surveys were randomly assigned to the training dataset and 305 to the validation dataset.

In the baseline homogeneity analysis, the training and the validation datasets were compared (table 1). In the training dataset, a significantly higher percentage of patients had some degree of recognised general disability (49.1 vs 39.7%, p=0.03); likewise, the patients in the training sample had been diagnosed at an earlier age (27.9 ± 10.4 vs 30 ± 11.5 years, *p* = 0.015) and had longer disease duration (14.2 ± 10.2 vs 12.8 ±9.9 years, *p* = 0.03). No other significant differences were found between the two datasets.

### Bivariate and multivariate analyses for work disability

To determine the factors predicting work disability, we analysed 544 surveys in the training dataset completed by respondents from the active working population. Fifty-three patients in this group had a recognised work disability. All the variables significantly related to the degree of disability in the bivariate analysis (supplementary table 2) were included in the multivariate logistic regression model.

The final model for predicting the granting of a work disability benefit included all the variables that were significant in the multivariate analysis including age, number of daily bowel movements, number of surgeries, number of medical examinations in the last year and body mass index (BMI). Three additional variables considered clinically important despite not being statistically significant –time of IBD evolution (p=0.2), fecal incontinence (p=0.73) and ostomy (p=0.12)– were also included (table 2). The AUROC of this model was 0.811 (Supplementary figure 1a).

**Table 2:** Multivariate analysis and modelling

| **Work disability** | **Odds Ratio** | **95% Wald**<br>**Confidence Limits** |  | **p** |
| --- | --- | --- | --- | --- |
| Age | *1.044 | 1.010 | 1.080 | 0.016 |
| Time of disease evolution ($\leq 5$ vs $\geq 6$) | 2.033 | 0.706 | 5.848 | 0.2 |
| Liquid fecal incontinence | 1.269 | 0.642 | 2.513 | 0.73 |
| Bowel movements per day ($>6$ vs $\leq 3$) | *3.922 | 1.631 | 9.434 | 0.008 |
| Bowel movements per day $>6$ vs 4-6) | *3.448 | 1.418 | 8.403 | |
| Surgical interventions due to IBD ($\geq 3$ vs 0) | *4.739 | 2.079 | 10.870 | 0.001 |
| Surgical interventions due to IBD ($\geq 3$ vs 1-2) | 1.447 | 0.674 | 3.106 | |
| Ostomy | 2.110 | 0.769 | 5.780 | 0.12 |
| Medical tests for IBD in the last year ($\geq 4$ vs $\leq 3$) | *2.427 | 1.087 | 5.435 | 0.04 |
| BMI ($<18.5$ vs $>18.5$) | *3.525 | 1.309 | 9.495 | 0.01 |
| **General disability** |  |  |  |  |
| Time of disease evolution ($\leq 5$ vs $\geq 6$) | *6.369 | 3.717 | 10.870 | 0.001 |
| Immunosuppressant treatment | *2.165 | 1.381 | 3.390 | 0.001 |
| Liquid fecal incontinence | 1.124 | 0.701 | 1.802 | 0.6 |
| Bowel movements per day ($>6$ vs $\leq 3$) | 1.372 | 0.737 | 2.551 | 0.6 |
| Bowel movements per day ($>6$ vs 4-6) | 1.272 | 0.662 | 2.445 | |
| Psychiatric disease | *2.618 | 1.590 | 4.310 | 0.001 |
| Surgical interventions due to IBD ($\geq 3$ vs 0) | *5.495 | 2.625 | 11.494 | 0.001 |
| Surgical interventions due to IBD ($\geq 3$ vs 1-2) | *2.809 | 1.271 | 6.211 | |
| Ostomy | *3.322 | 1.067 | 10.309 | 0.03 |
| Arthralgia or arthritis | *1.558 | 1.040 | 2.331 | 0.03 |
| **Severity of general disability** |  |  |  |  |
| Age | *1.023 | 1.000 | 1.046 | 0.06 |
| Time of disease evolution ($\leq 5$ vs $\geq 6$) | *3.676 | 1.866 | 7.246 | 0.001 |
| Liquid fecal incontinence | 1.244 | 0.766 | 2.019 | 0.25 |
| Bowel movements per day ($>6$ vs $\leq 3$) | 1.704 | 0.853 | 3.401 | 0.1 |
| Bowel movements per day ($>6$ vs 4-6) | *2.257 | 1.094 | 4.651 | |
| Psychiatric disease | *1.776 | 1.092 | 2.882 | 0.02 |
| Surgical interventions due to IBD ($\geq 3$ vs 0) | *4.016 | 2.066 | 7.813 | 0.001 |
| Surgical interventions due to IBD ($\geq 3$ vs 1-2) | *2.326 | 1.183 | 4.566 | |
| Ostomy | 2.110 | 0.922 | 4.831 | 0.16 |
| Arthralgia or arthritis | *1.629 | 1.026 | 2.584 | 0.4 |

### Bivariate and multivariate analyses for general disability

Six hundred and eighteen patients were eligible for the analysis. Seven patients who had submitted a request for disability benefit and were awaiting a response were excluded. Three hundred and seven patients were recognised as having some degree of general disability.

All the variables significantly related to the degree of disability in the bivariate analysis (supplementary table 2) were included in the multivariate logistic regression model.

The final model for predicting the granting of a degree of general disability included all the variables that were significant in the multivariate analysis –IBD time of evolution, immunosuppressant treatment, psychiatric comorbidity, number of surgeries, ostomy and arthropathy– plus two non-significant variables considered clinically important: incontinence (p=0.6) and number of bowel movements per day (p=0.6) (table 2). The AUROC was 0.806 (suppl. figure 1b).

### Bivariate and multivariate analyses of the degree of general disability

All the variables significantly related to the degree of disability in the bivariate analysis (supplementary table 2) were included in the multivariate ordinal logistic regression model. The final model for predicting the degree of general disability included all the variables that were significant in the multivariate analysis –age, time of evolution of the disease, psychiatric comorbidity, number of surgeries, ostomy or arthropathy– plus incontinence (0.25), number of movements per day (p=0.1) and ostomy (p=0.16) (table 2). The AUROC was 0.629.

### Score modelling

Eight variables were considered eligible for inclusion in the score: time of evolution of the disease, psychiatric comorbidities, arthropathy, and the number of surgeries, which were all significantly related to disability in at least two of the multivariate analyses. Ostomy, mean number of bowel movements per day and age were also included because they were significant in one of the three multivariate logistic regression models and were considered clinically significant. Finally, though not significant in any of the three multivariate analyses, fecal incontinence was included due to its major impact on patients’ quality of life and disability as recorded in the literature [32].

Initially, two different models were designed (table 3). In the first, each variable scored one point (unweighted IBD objective disability Index, U-IBDODI). In the second model (weighted IBD objective disability index, W-IBDODI) each variable had a different weight calculated according to the average of their Odds Ratio on the three logistic regressions.

**Table 3:** IBDODI models

|  |  | **W-IBDODI** | **U-IBDODI** |
| --- | --- | --- | --- |
| Age | > 50 years | 1.3 | 1 |
| Time of disease evolution | > 5 years | 4 | 1 |
| Liquid fecal incontinence | Yes | 1 | 1 |
| Bowel movements per day | > 6 | 2.2 | 1 |
| Psychiatric comorbidity | YES | 2.1 | 1 |
| Surgeries | 0 | 0 | 0 |
|  | 1-2 | 2,5 | 0 |
|  | >2 | 5 | 1 |
| Ostomy | Yes | 5 | 1 |
| Arthritis | Yes | 1.6 | 1 |

Possible U-IBDODI values ranged from zero to eight. In the training dataset, patients scored a minimum value of zero and a maximum value of seven. Mean and median values were 2.4 ± 1.4 and 2 respectively (suppl. Figure 2a). Possible W-IBDODI values ranged from zero to 19.4. In the training set, patients’ minimum value was zero and the maximum value 17.2. Mean and median values were 7.3 ± 3.9 and 7.2 respectively (suppl. Figure 2b).

In the training dataset, both U-IBDODI and W-IBDODI were able to distinguish between patients with and without a work disability benefit. Respective mean values were 3.7 ± 1.3 and 10.9 ± 3.7 for patients with work disability and 2.3 ± 1.4 and 6.9 ± 3.7 for patients without. Both *p* values were <0.001 (table 4). The respective AUROCs were 0.76 and 0.771 (Supplementary figure 3, a and b).

**Table 4:** U-IBDODI and W-IBDODI values for patients with/without work or general disability in training and validation datasets

|  | Training dataset |  | Validation dataset |  |
| --- | --- | --- | --- | --- |
|  |  | p |  | p |
| **Work disability** |  |  |  |  |
| U-IBDODI |  | 0.001 |  | 0.001 |
| • With disability | 3.72 ±1.34 |  | 4.6 ±1.6 |  |
| • Without disability | 2.32 ±1.38 |  | 2.4 ±1.5 |  |
| W-IBDODI |  | 0.001 |  | 0.001 |
| • With disability | 10.9 ±3.7 |  | 11.2 ±3.3 |  |
| • Without disability | 7 ±3.7 |  | 6.3 ±3.5 |  |
| **General disability** |  |  |  |  |
| U-IBDODI |  | 0.001 |  | 0.001 |
| • With disability | 3 ±1.4 |  | 3.2 ±1.8 |  |
| • Without disability | 1.9 ±1.3 |  | 2.1 ±1.5 |  |
| W-IBDODI |  | 0.001 |  | 0.001 |
| • With disability | 9.2 ±3.5 |  | 8.1 ±4 |  |
| • Without disability | 5.6 ±3.3 |  | 5.6 ±3.6 |  |
| **Degree of general disability** |  |  |  |  |
| U-IBDODI |  | 0.001 |  | 0.001 |
| • 0 (no disability) | 2.39 ±1.44 |  | 2.71 ±1.40 |  |
| • 1-32 | 2.28 ± 1.18 |  | 2.04 ±1.30 |  |
| • 33-54 | 3.01 ±1.27 |  | 3.24 ±1.66 |  |
| • 55+ | 4.24 ±1.24 |  | 3.94 ±1.95 |  |
| W-IBDODI |  | 0.001 |  | 0.001 |
| • 0 (no disability) | 6.5 ±4.04 |  | 7.68 ±3.31 |  |
| • 1-32 | 7.58 ±3.06 |  | 5.73 ±2.94 |  |
| • 33-54 | 9.15 ±3.42 |  | 8.35 ±3.96 |  |
| • 55+ | 12.53 ±2.73 |  | 9.16 ±4.14 |  |

The mean values of U-IBDODI and W-IBDODI for general disability are shown in table 4. U-IBDODI had an AUROC of 0.713 and W-IBDODI one of 0.765 (Supplementary figures 3 c and d). U-IBDODI mean values ranged from 2.28 in patients with mild disability to 4.24 in those with severe disability (table 4); the respective values for W-IBDODI were 7.6 and 12.5 (table 4). AUROCs for the discrimination of the degree of general disability were 0.669 for U-IBDODI and 0.686 for W-IBDODI; the two scores achieved significant differentiation only between groups with high (>55%) or low (<33%) percentages of general disability (Supplementary table 3).

### Score validation

In the validation dataset the median U-IBDODI score was 2.61 ± 1.66, ranging from zero to eight (Figure 2a), and the W-IBDODI score ranged from zero to 17.2. Mean and median W-IBDODI values were 6.79 ± 3.9 and 6.55 respectively (Figure 2b).

**Figure 2:**
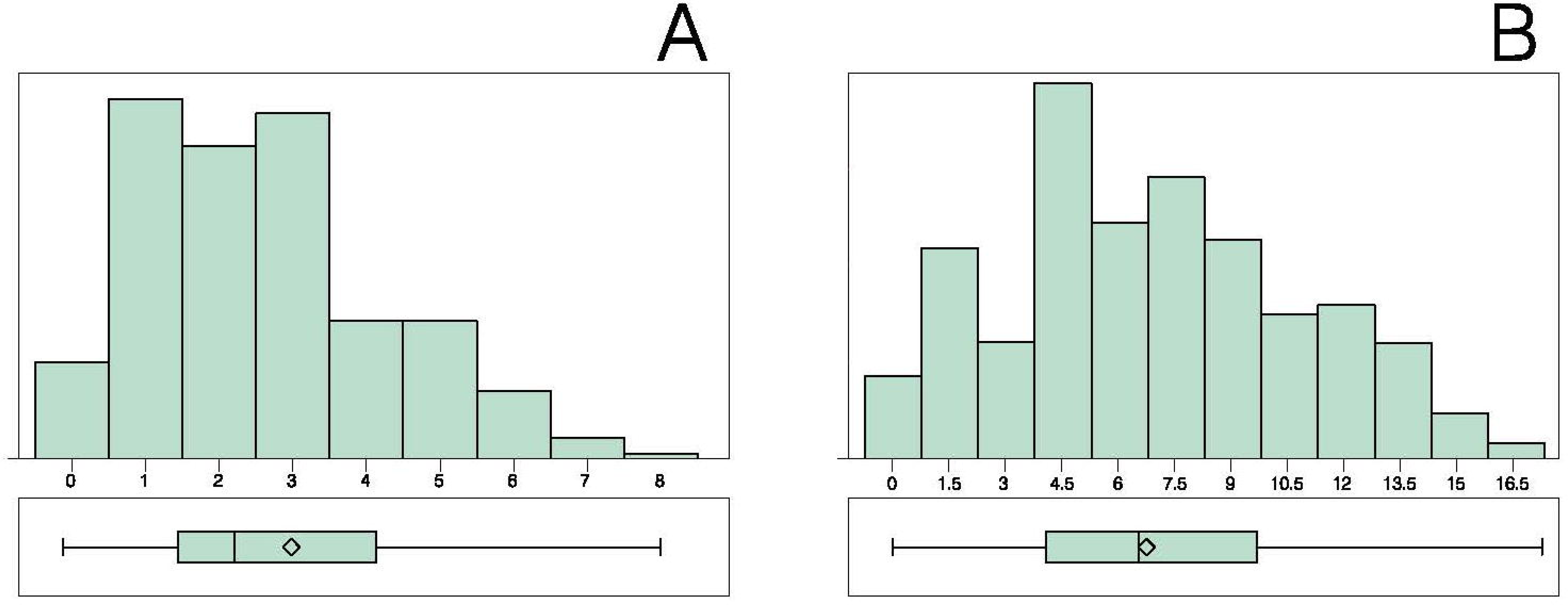
The distribution of U-IBDODI (a) and W-IBDODI (b) values in the validation dataset

Mean U-IBDODI and W-IBDODI scores in the validation dataset are shown in table 4 for both general and work disability. The AUROCs of U-IBDODI and W-IBDODI were 0.839 and 0.837 respectively for work disability (Fig. 3 a-b), and 0.675 and 0.676 respectively for general disability (Figs 3 c-d).

**Figure 3:**
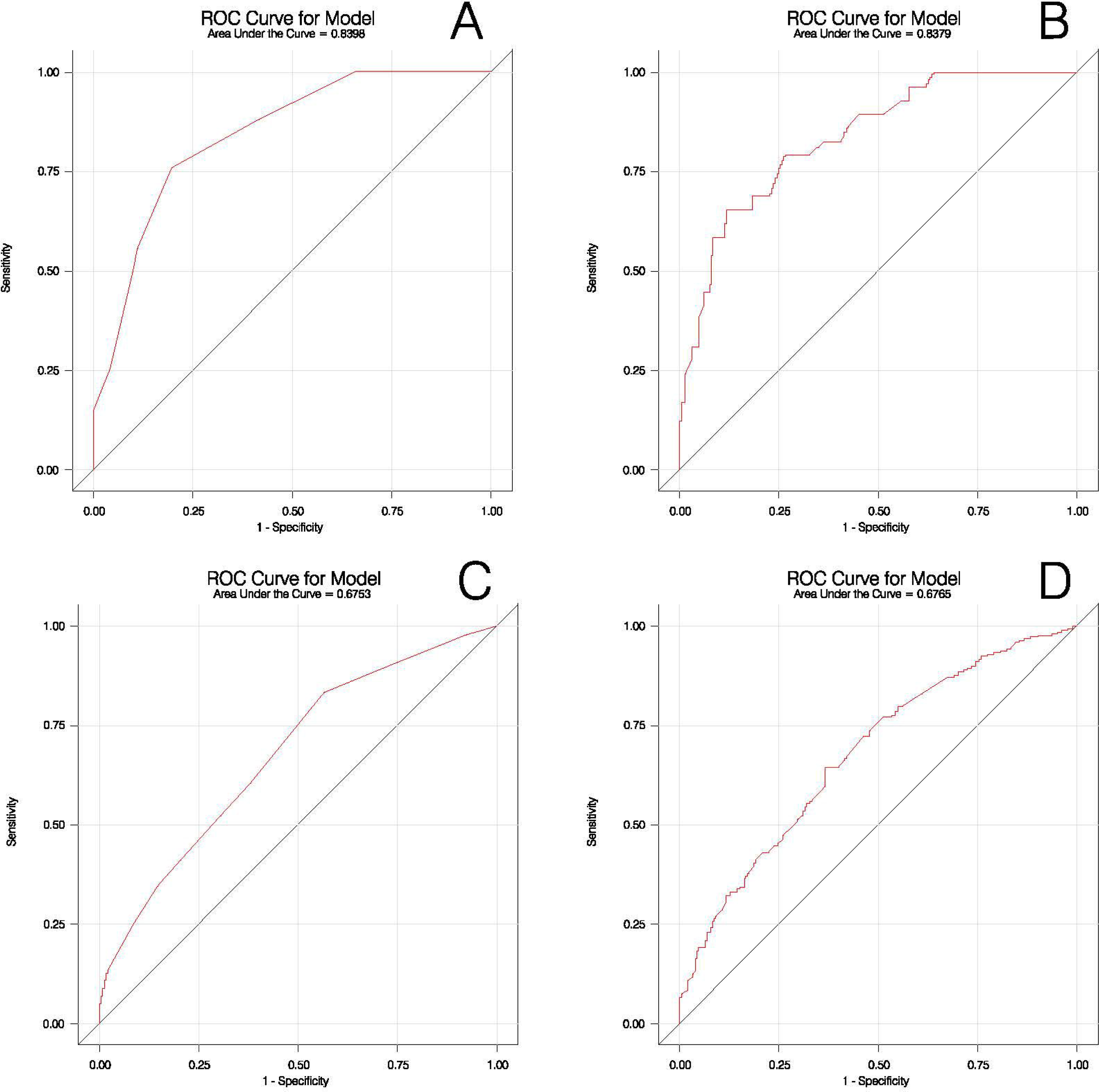
U-IBDODI and W-IBDODI AUROCs for work disability (a-b) and for general disability (c-d) in the validation dataset

U-IBDODI and W-IBDODI values for degree of general disability are shown in table 4. The AUROCs were 0.642 for U-IBDODI and 0.606 for W-IBDODI (Supplementary table 3).

## Discussion

To our knowledge, the present study is the first to develop and validate a work disability index based exclusively on variables that can be objectively measured. The IBDODI is able to accurately predict long-term disability, with an AUROC for work disability above 0.8. The AUROCs in the training and validation sets were similar. This index also presents advantages over previous scoring systems, as it is based only on objective and measurable values, a feature that makes it useful for guiding administrative decisions regarding the recognition of work disability and the award of work disability grants. The IBDODI can also be used as an online self-reported index. The fact that it includes only eight items facilitates its application and renders it useful for large surveys or epidemiological studies. The two scores designed – the U-IBDODI and the W-IBDODI – were equally valid for measuring disability in patients with IBD. However, since the U-IBDODI is easier to calculate than the W-IBDODI and achieves a similar level of accuracy, it is probably the preferred option for objectively measuring work disability.

The IBDODI was also able to predict general disability, although in this case the score seems less reliable. This poorer performance may be due to limitations of the score itself, or may reflect a greater heterogeneity in the administrative evaluation of general disability.

IBD-related disability has received growing attention in recent times. There have been many different attempts to create a disability index [5,13,16]. Feagan et al. proposed disability as an endpoint in therapeutic trials [12] and the STRIDE II consensus [33] included the absence of disability as a long-term target in IBD patient management. However, measuring disability is a challenge. Acute flares of the disease may be extremely disabling; however, this acute and short-term disability is generally reversible and is not an accurate predictor of long-term limitations related to IBD. Furthermore, the items measured in assessments of short-term disability related to disease activity are similar to those measured by IBD quality of life (QoL) indexes. Therefore, it is important to establish clearly what kind of disability we want to measure. To avoid overlapping with QoL measures and with acute flare symptoms, in our opinion, disability measures should incorporate chronicity and the issue of reversibility. For example, disease activity should only be considered as chronically disabling in the context of persistent activity with multiple treatment failures that render adequate future medical or surgical adequate control unlikely. However, symptoms due to structural bowel lesions or surgical sequelae should be considered as consistent markers of disability. Thus, the evaluation of the irreversibility of the clinical situation causing functional impairment should be incorporated into any score devised to evaluate long-term disability [34]. Previous disability scores have not taken into account the irreversibility of functional impairment (or have done so only partially), thus limiting their ability to measure permanent disability [14,16,17].

In three different logistic regressions we identified a number of variables that were consistently related with administratively recognised chronic disability: age over 50, disease evolution longer than 6 years the increase in the number of stools, previous surgical treatments, ostomy and comorbidities such as arthralgia or psychiatric illness [5,16]. Many of these items have been included in previous disability scores: for example, the number of bowel movements per day and extra-intestinal symptoms such as arthralgia or psychiatric diseases appear in the IBD-DI, the UCWDQ and the CDWDQ [14,16,17]. In this regard, we incorporated fecal incontinence (a variable that did not achieve significance in our models) because it has been consistently included in all previous disability scores [16,17]. The IBDODI also includes variables such as the patient’s age, time since IBD diagnosis, number of surgical interventions and the presence of an ostomy which were not included in other disability scores but which were associated with the presence of disability in our multivariate analysis and in previous observational studies [5,22]. In particular, ostomy was included because a previous study by our group showed it to be one of the strongest predictors of having a recognised work disability [22].

The study has a number of limitations, some inherent to all studies trying to evaluate disability and others related to the study design. One major limitation is the lack of a reliable standard to measure disability. Even the most widely accepted classification, the International Classification of Functioning, Disability, and Health (ICF) of the World Health Organization (WHO) [35] included multiple subjective indicators of disability such as the capacity for concentration on a task, the ability to find solutions in daily life or having to work at a lower intensity due to disease. For this reason, despite their evident limitations, we considered the results of the administrative evaluations of the Social Security authorities for granting disability benefits as the standard for designing and validating the scores. Although this evaluation is heterogeneous [22], it was, in our opinion, the most reliable measurement available.

Another limitation of our study is that the data come from a survey that was publicised through the patients’ association website. This means that subjects who do not have access to the Internet or who do not participate in patients’ associations were excluded. As a consequence, the questionnaires analysed probably include an above-average proportion of young and more severely ill patients and may not be representative of the whole IBD population. A further limitation is that the survey was conducted in a single country (Spain). The validation of translations of other indices of quality of life or general and work disability has always shown identical performance when English-language questionnaires have been translated into Spanish [29,36]; however, the validation of the questionnaire in other languages and populations is mandatory before the widespread use of the index can be recommended.

Finally, although our questionnaire was designed to establish a set of objective variables related to disability, these variables were obtained from a self-reported questionnaire. This fact is both a strength and a limitation. The questionnaire seems useful as a self-reported online tool for detecting work disability; however, in the setting of the administrative evaluation of disability in an individual patient, the score needs to be validated in medical or administrative records to ensure the absence of bias in the reporting of the data.

A major strength of the study is the large number of IBD patients included, a feature that increases the reliability of the analysis. In addition, as there was no relation to any benefit for patients, the risk of bias in reporting the variables is low.

Further developments of the IBDODI should include its validation in other languages and in different populations. In the meantime, the results are already available for use. It is reasonable to suggest that medical reports for requesting the recognition of a disability should include a description of the variables detected as independent predictors of disability in the present study. It is also reasonable for evaluators to request and analyse these parameters so to guide administrative decisions.

In conclusion, the IBDODI appears to be a reliable tool for assessing long-term disability in IBD especially long-term work disability. It can help health authorities to objectively evaluate disability and thus increase levels of impartiality and fairness in administrative decisions. Furthermore, this simple, easy-to-use tool allows the assessment of disability due to IBD in large-scale surveys.

## Data Availability

All data produced in the present work are contained in the manuscript

## Supplementary figure legend

**Supplementary figure 1:**
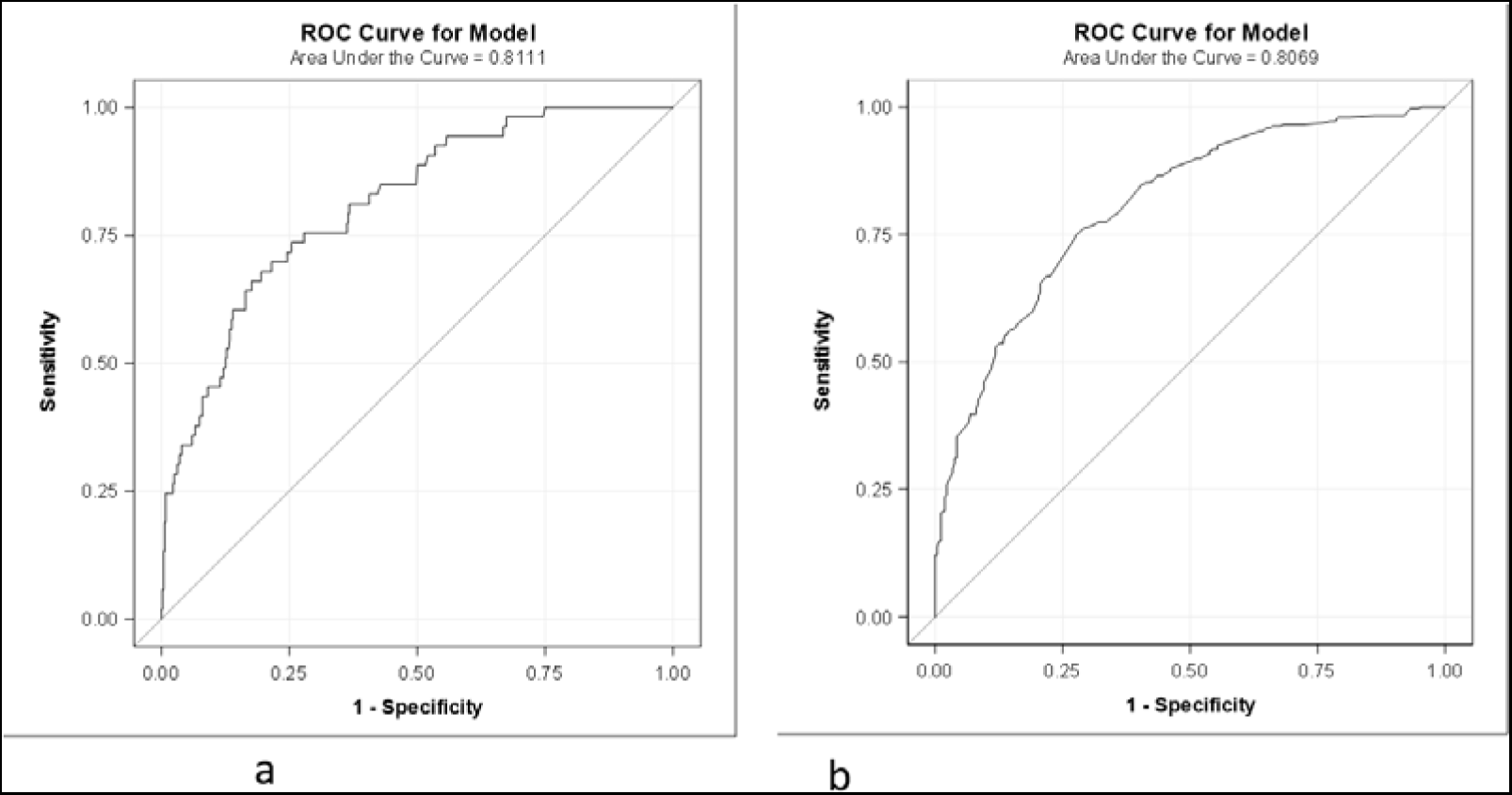
AUROC of the model for predicting the granting of work disability (a), general disability (b) with all the variables significant in the work disability logistic regression model in the training set.

**Supplementary figure 2:**
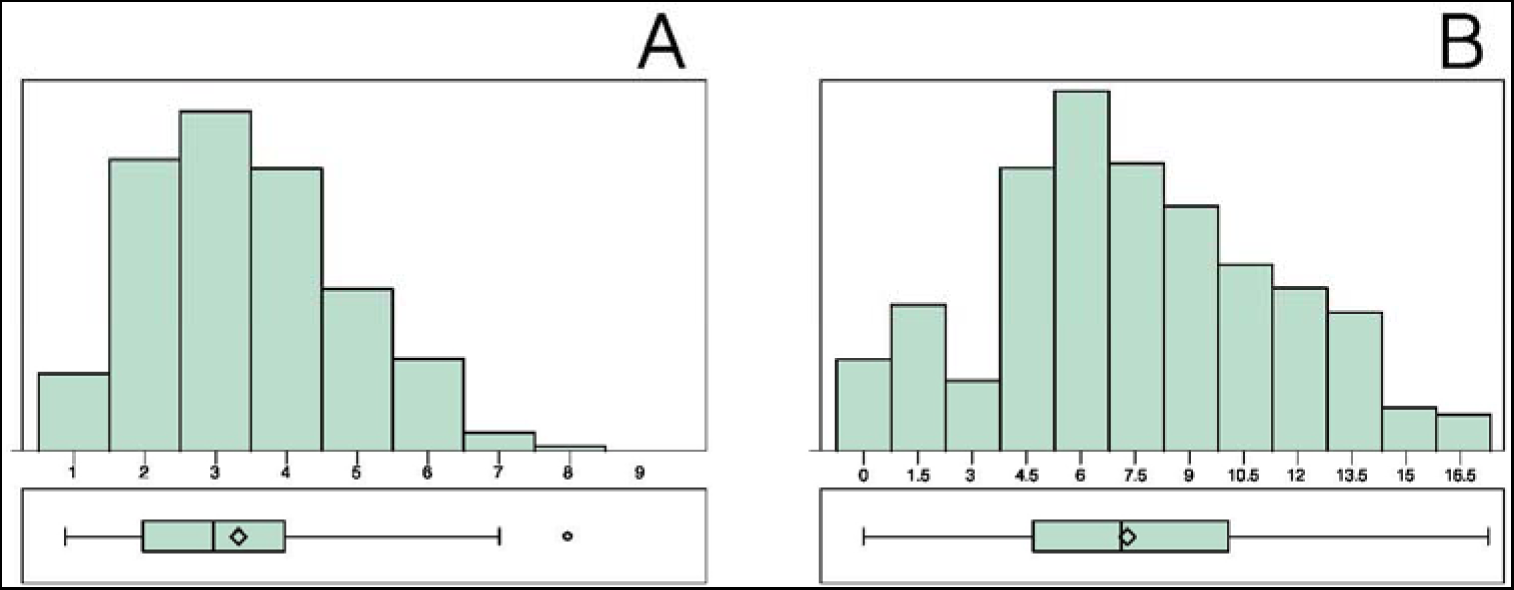
Distribution of the U-IBDODI (a) and W-IBDODI (b) training dataset results.

**Supplementary figure 3:**
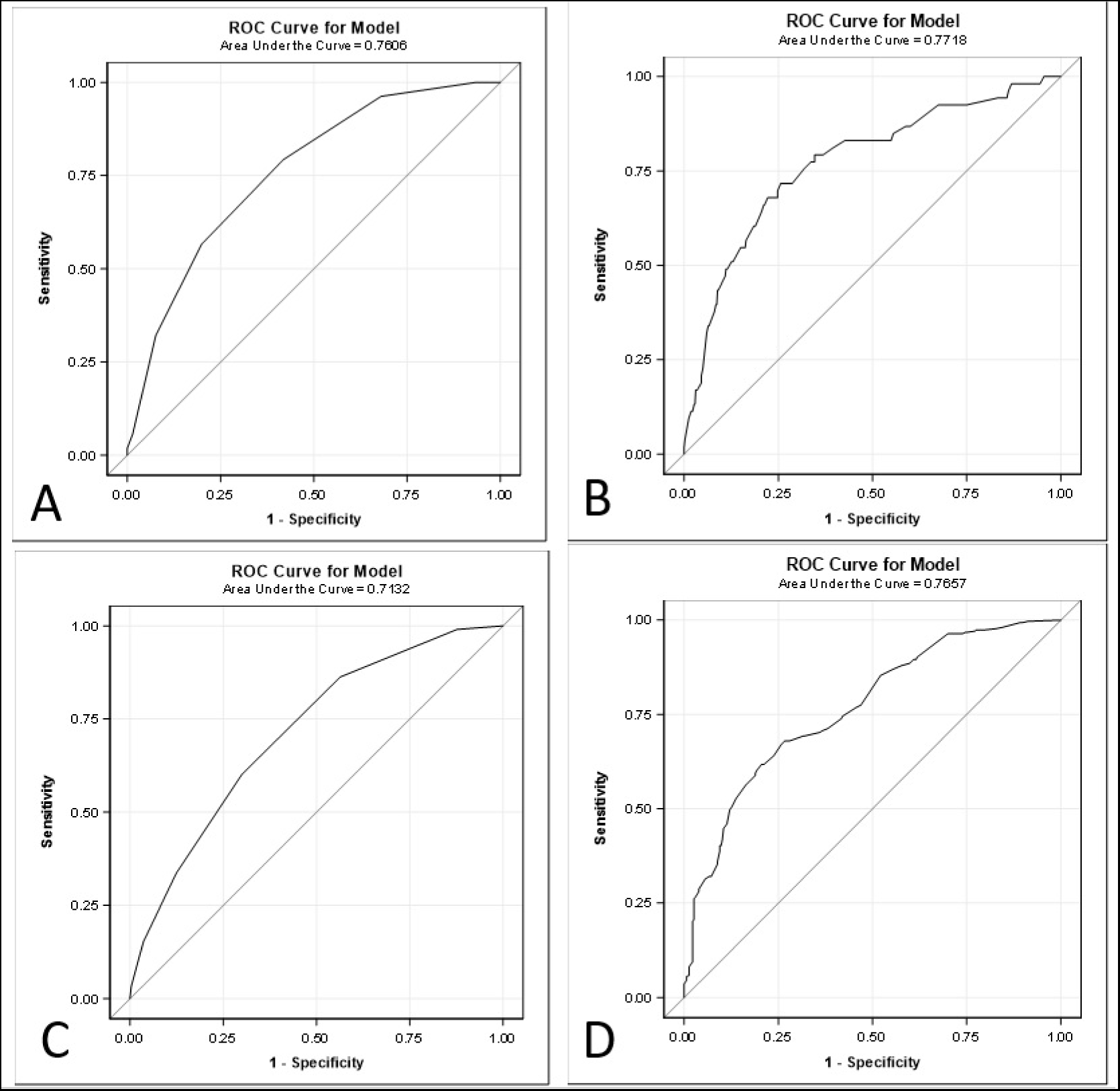
AUROC curves in the U-IBDODI and W-IBDODI training datasets for work disability (a-b) and for general disability (c-d)

## SUPPLEMENTARY TABLE LEGEND

**Supplementary table 1:** TRIPOD Development-and-Validation Checklist

| Section/Topic | Item |  | Checklist Item | Page |
| --- | --- | --- | --- | --- |
| **Title and abstract** |  |  |  |  |
| Title | 1 | D;V | Identify the study as developing and/or validating a multivariable prediction model, the target population, and the outcome to be predicted. | 1 |
| Abstract | 2 | D;V | Provide a summary of objectives, study design, setting, participants, sample size, predictors, outcome, statistical analysis, results, and conclusions. | 2 |
| **Introduction** |  |  |  |  |
| Background and objectives | 3a | D;V | Explain the medical context (including whether diagnostic or prognostic) and rationale for developing or validating the multivariable prediction model, including references to existing models. | 4 |
|  | 3b | D;V | Specify the objectives, including whether the study describes the development or validation of the model or both. | 5 |
| **Methods** |  |  |  |  |
| Source of data | 4a | D;V | Describe the study design or source of data (e.g., randomized trial, cohort, or registry data), separately for the development and validation data sets, if applicable. | 6 |
|  | 4b | D;V | Specify the key study dates, including start of accrual; end of accrual; and, if applicable, end of follow-up. | 6 |
| Participants | 5a | D;V | Specify key elements of the study setting (e.g., primary care, secondary care, general population) including number and location of centers. | 6 |
|  | 5b | D;V | Describe eligibility criteria for participants. | 6 |
|  | 5c | D;V | Give details of treatments received, if relevant. | n/a |
| Outcome | 6a | D;V | Clearly define the outcome that is predicted by the prediction model, including how and when assessed. | 7 |
|  | 6b | D;V | Report any actions to blind assessment of the outcome to be predicted. | 6 |
| Predictors | 7a | D;V | Clearly define all predictors used in developing or validating the multivariable prediction model, including how and when they were measured. | 6-7 |
|  | 7b | D;V | Report any actions to blind assessment of predictors for the outcome and other predictors. | n/a |
| Sample size | 8 | D;V | Explain how the study size was arrived at. | 9 |
| Missing data | 9 | D;V | Describe how missing data were handled (e.g., complete-case analysis, single imputation, multiple imputation) with details of any imputation method. | 6-23 |
| Statistical analysis methods | 10a | D | Describe how predictors were handled in the analyses. | 8-9 |
|  | 10b | D | Specify type of model, all model-building procedures (including any predictor selection), and method for internal validation. | 8-9 |
|  | 10c | V | For validation, describe how the predictions were calculated. | 9 |
|  | 10d | D;V | Specify all measures used to assess model performance and, if relevant, to compare multiple models. | 9 |
|  | 10e | V | Describe any model updating (e.g., recalibration) arising from the validation, if done. | n/a |
| Risk groups | 11 | D;V | Provide details on how risk groups were created, if done. | n/a |
| Development vs. validation | 12 | V | For validation, identify any differences from the development data in setting, eligibility criteria, outcome, and predictors. | Table 1 |
| **Results** |  |  |  |  |
| Participants | 13a | D;V | Describe the flow of participants through the study, including the number of participants with and without the outcome and, if applicable, a summary of the follow-up time. A diagram may be helpful. | Figure 1 |
|  | 13b | D;V | Describe the characteristics of the participants (basic demographics, clinical features, available predictors), including the number of participants with missing data for predictors and outcome. | Table 1 |
|  | 13c | V | For validation, show a comparison with the development data of the distribution of important variables (demographics, predictors and outcome). | Table 1 |
| Model development | 14a | D | Specify the number of participants and outcome events in each analysis. | 11 |
|  | 14b | D | If done, report the unadjusted association between each candidate predictor and outcome. | Supplementary table 2 |
| Model specification | 15a | D | Present the full prediction model to allow predictions for individuals (i.e., all regression coefficients, and model intercept or baseline survival at a given time point). | Table 2 |
|  | 15b | D | Explain how to use the prediction model. | 13-14 |
| Model performance | 16 | D;V | Report performance measures (with CIs) for the prediction model. | 14 + figure 3 |
| Model-updating | 17 | V | If done, report the results from any model updating (i.e., model specification, model performance). | n/a |
| **Discussion** |  |  |  |  |
| Limitations | 18 | D;V | Discuss any limitations of the study (such as nonrepresentative sample, few events per predictor, missing data). | 16 |
| Interpretation | 19a | V | For validation, discuss the results with reference to performance in the development data, and any other validation data. | 15 |
|  | 19b | D;V | Give an overall interpretation of the results, considering objectives, limitations, results from similar studies, and other relevant evidence. | Discussion |
| Implications | 20 | D;V | Discuss the potential clinical use of the model and implications for future research. | 17 |
| **Other information** |  |  |  |  |
| Supplementary information | 21 | D;V | Provide information about the availability of supplementary resources, such as study protocol, Web calculator, and data sets. | n/a |
| Funding | 22 | D;V | Give the source of funding and the role of the funders for the present study. | 3 |

**Supplementary table 2:** Bivariate analysis for work and general disability and for the degree of general disability

|  | Work disability |  | General disability |  | Degree of general disability |  |
| --- | --- | --- | --- | --- | --- | --- |
| *Variable* | *P-Value* |  | *P-Value* |  | *P-Value* |  |
| Age | 0.002 | * | <0.001 | * | <0.001 | * |
| Biological sex | 0.526 |  | 0.316 |  | 0.520 |  |
| Region of residence | 0.211 |  | 0.001 | * | <0.001 | * |
| UC vs CD | 0.055 |  | <0.001 | * | 0.805 |  |
| Year of diagnosis | 0.002 | * | <0.001 | * | <0.001 | * |
| Age at time of diagnosis | 0.832 |  | 0.012 | * | 0.744 |  |
| Time of disease evolution | 0.002 | * | <0.001 | * | <0.001 | * |
| Pharmacological treatment | 0.214 |  | <0.001 | * | 0.453 |  |
| Immunosuppressant treatment | 0.044 | * | <0.001 | * | 0.447 |  |
| Corticosteroids | 0.822 |  | 0.250 |  | 0.135 |  |
| Type of biological treatment | 0.003 | * | 0.646 |  | 0.262 |  |
| Solid incontinence | 0.120 |  | 0.002 | * | 0.612 |  |
| Liquid incontinenes | <0.001 | * | <0.001 | * | 0.001 | * |
| Gas incontinence | 0.105 |  | 0.393 |  | 0.289 |  |
| Incontinence (Wexner index) | 0.005 | * | <0.001 | * | 0.275 |  |
| Incontinence | 0.012 | * | 0.002 | * | 0.295 |  |
| Bowel movements per day | <0.001 | * | 0.036 | * | 0.023 | * |
| UC: extension | 0.369 |  | 0.300 |  | 0.665 |  |
| UC: Bowel frequency in last week | 0.017 | * | 0.024 | * | 0.540 |  |
| UC: Depositional urgency | 0.874 |  | 0.055 |  | 0.042 | * |
| UC: Rectal bleeding | 0.262 |  | 0.685 |  | 0.458 |  |
| UC: Global assessment | 0.109 |  | 0.133 |  | 0.330 |  |
| UC: Extraintestinal manifestation (EIM) arthritis | 0.909 |  | 0.273 |  | 0.261 |  |
| EIM (UC): ocular manifestation (uveitis, episcleritis, scleritis) | 0.892 |  | 0.016 | * | 0.437 |  |
| EIM (UC) erythema nodosum | 1.000 |  | 0.098 |  | 0.505 |  |
| EIM (UC) pyoderma gangrenosum | 0.188 |  | 0.026 | * | 0.618 |  |
| No EIM (UC) | 0.576 |  | 0.114 |  | 0.413 |  |
| Number of EIM (UC) | 0.553 |  | 0.143 |  | 0.490 |  |
| SCCAI: Simple clinical colitis activity index | 0.086 |  | 0.006 | * | 0.166 |  |
| SCCAI active vs inactive disease | 0.244 |  | 0.282 |  | 0.171 |  |
| Grouped EIM | 0.047 | * | 0.007 | * | 0.491 |  |
| CD: Localisation | 0.178 |  | 0.048 | * | 0.131 |  |
| CD: Behaviour | 0.031 | * | 0.038 | * | 0.101 |  |
| CD: Perianal disease | 0.101 |  | <0.001 | * | 0.028 | * |
| CD: Harvey Bradshaw index | <0.001 | * | 0.002 | * | 0.121 |  |
| CD: Global assessment in last week | 0.011 | * | 0.240 |  | 0.355 |  |
| CD: Abdominal pain in last week | 0.247 |  | 0.018 | * | 0.017 | * |
| CD: Number of liquid/soft stools | <0.001 | * | <0.001 | * | 0.013 | * |
| CD: Abdominal mass | 0.247 |  | 0.107 |  | 0.325 |  |
| EIM (CD) arthritis | 0.019 | * | 0.012 | * | 0.121 |  |
| EIM (CD) uveitis | 0.769 |  | 0.092 |  | 0.069 |  |
| EIM (CD) erythema nodosum | 0.710 |  | 0.053 |  | 0.145 |  |
| EIM (CD) aphthous ulcers | 0.135 |  | 0.437 |  | 0.463 |  |
| EIM (CD) pyoderma gangrenosum | 0.465 |  | 0.633 |  | 0.314 |  |
| EIM (CD) anal fissure | 0.043 | * | 0.399 |  | 0.012 | * |
| EIM (CD) abscess | <0.001 | * | 0.168 |  | 0.057 |  |
| Perianal fistula | 0.756 |  | 0.336 |  | 0.544 |  |
| New fistula | 0.832 |  | 0.540 |  | 0.815 |  |
| No EIM | 0.198 |  | 0.178 |  | 0.301 |  |
| CD: number of EIM | 0.058 |  | 0.364 |  | 0.536 |  |
| Use of psychiatric drugs | 0.056 |  | <0.001 | * | 0.723 |  |
| Psychiatric disease | 0.003 | * | <0.001 | * | 0.014 | * |
| Medications and/or diagnosed disease | 0.011 | * | <0.001 | * | 0.125 |  |
| Type of psychiatric disease | 0.637 |  | 0.416 |  | 0.451 |  |
| Surgery due to IBD (abdominal and perianal) | <0.001 | * | <0.001 | * | <0.001 | * |
| Number of surgeries due to IBD (abdominal and perianal) | <0.001 | * | <0.001 | * | <0.001 | * |
| Abdominal surgery due to IBD | <0.001 | * | <0.001 | * | <0.001 | * |
| Number of abdominal surgeries due to IBD | <0.001 | * | <0.001 | * | <0.001 | * |
| Intestinal resections | <0.001 | * | <0.001 | * | <0.001 | * |
| Number of intestinal resections | <0.001 | * | <0.001 | * | <0.001 | * |
| Surgery due to perianal disease | <0.001 | * | <0.001 | * | <0.001 | * |
| Number of surgeries due to perianal disease | <0.001 | * | <0.001 | * | 0.002 | * |
| Ostomy | 0.001 | * | <0.001 | * | <0.001 | * |
| Ileoanal reservoir | 0.047 | * | 0.103 |  | 0.246 |  |
| Ostomy and ileoanal reservoir | <0.001 | * | <0.001 | * | <0.001 | * |
| Days of sick leave in the last year due to IBD | <0.001 | * | <0.001 | * | 0.433 |  |
| Specialised medical visits for IBD in the last year | 0.031 | * | 0.007 | * | 0.546 |  |
| Days of hospitalisation for IBD in the last year | 0.009 | * | 0.543 |  | 0.001 | * |
| Medical tests for IBD in the last year | 0.012 | * | 0.014 | * | 0.563 |  |
| BMI | 0.342 |  | 0.125 |  | 0.539 |  |
| BMI categorised | 0.011 | * | 0.183 |  | 0.300 |  |
| Blood in stool | 0.752 |  | 0.394 |  | 0.173 |  |
| Arthralgias or arthritis | 0.009 | * | <0.001 | * | 0.017 | * |

**Supplementary table 3:** AUROCs of U-IBDODI and W-IBDODI for the degree of general disability

|  |  | U-IBDODI training dataset<br>AUC | W-IBDODI training dataset<br>AUC | U-IBDODI validation dataset<br>AUC | W-IBDODI validation dataset<br>AUC |
| --- | --- | --- | --- | --- | --- |
| 0 | 1-32 | 0.5000 | 0.5064 | 0.5000 | 0.5000 |
| 0 | 33-54 | 0.6150 | 0.6875 | 0.5943 | 0.5559 |
| 0 | 55+ | 0.8238 | 0.8846 | 0.6887 | 0.6048 |
| 1-32 | 33-54 | 0.6544 | 0.6178 | 0.7356 | 0.7136 |
| 1-32 | 55+ | 0.8681 | 0.8851 | 0.8056 | 0.7558 |
| 33-54 | 55+ | 0.5543 | 0.5401 | 0.5334 | 0.5084 |

